# Longitudinal Placental Blood Volume Measurements in Zika-Infected Rhesus Macaques Using Ferumoxytol Enhanced MRI

**DOI:** 10.1101/2025.03.27.25323585

**Authors:** Ruiming Chen, Sydney Nguyen, Megan E. Murphy, Kathleen M. Antony, Sean B. Fain, Dinesh Shah, Thaddeus Golos, Oliver Wieben, Kevin M. Johnson

## Abstract

**Introduction:** **Measures of** maternal fractional blood volume (mFBV) in the placenta holds potential to diagnose placental vasculature deficiencies. However, methods for quantitative mapping of blood volume are challenging to implement for clinical placenta evaluation. As a preliminary step towards human applications, this study assesses the feasibility of blood volume measurements using ferumoxytol enhanced variable flip angle (VFA) T1-mapping in Zika-infected rhesus macaques.

**Methods:** Seven pregnant rhesus macaques were imaged longitudinally at up to 3 timepoints across gestation (days 64.5±1.9, 100.8±3.9, and 145.3±1.8), corresponding to first, second, and third pregnancy trimester of the rhesus. Four animals received a Zika virus (ZIKV) injection into the amniotic fluid, while three control rhesus macaques received a saline injection. T1-weighted spoiled gradient echo sequences at four flip angles (2°, 6°, 10°, 14°) were used for quantitative mFBV assessment derived from pre- and post-contrast T1 mapping using ferumoxytol. Image quality assessment and segmentation assessment was performed on the full 3D coverage. Placental histopathology for all animals was analyzed by a professional pathologist with over 15 years of experience.

**Results:** All scans were successfully acquired and analyzed with no significant motion artifacts. 3D mFBV maps show regional heterogeneities within slices. FBV and total placental blood volume has an increasing trend with gestation.

**Discussion:** This study shows feasibilities to measure mFBV in non-human primates using ferumoxytol enhanced VFA T1-mapping.

## Introduction

Adequate development of utero-placental vasculature plays a critical role in placental health and fetal growth. Abnormal adaptation of placental vasculature to fetal demands can lead to mal-perfusion and gestational complications such as fetal growth restriction and preeclampsia ([1–3]). Therefore, there are potential benefits to accurately and noninvasively track and quantify placenta vascular development throughout the multiple stages of gestation, including early gestation where there is a potential window for interventions and treatments.

Clinically, ultrasound is widely used for imaging in pregnancy. The advantages of ultrasound include cost-effectiveness, accessibility, and imaging with high frame rates. However, ultrasound has limitations in assessing placental vascular health including relatively unreliable measurements of macroscopic placental blood flow to the placenta due to vessel tortuosity and lack of relevant quantitative imaging markers for regional functional assessment [4]. Meanwhile, MRI enables he interrogation of vascular function and health through measures such as macrovascular blood flow [5], global and local perfusion [6], oxygenation [7], and inflammation [8] of organs including the brain, heart, liver, and kidneys. Recent advances in placental MRI have enabled the assessment of quantitative markers such as placental perfusion [9], blood oxygenation level dependent (BOLD) effects [10], native R2*/T2* quantification [11], and direct blood flow measures [12]. These quantitative metrics provide unique depictions of placental health; however, they are often confounded by the placental anatomy and physiology. For example, T2* relaxometry is sensitive to oxygenation, but also to vessel architecture and the presence of iron and calcium. Further, vascular metrics from non-contrast perfusion MRI using arterial spin labeling (ASL) can be limited by the long transit time (10s of s) in the intervillous space and unique architecture of the spiral arteries, leading to underestimation of actual placental perfusion [9].

The use of a contrast agent can provide additional information regarding placental intervillous architecture, organization, flow and perfusion (9). Clinically, dynamic imaging of intravenously injected Gadolinium (Gd) based agents is routinely used to derive blood flow and blood volume measures across multiples organs. While Gadolinium has been used in pregnant non-human primate experimental models [13], and a pilot study in human pregnancy (n=6)[14], concerns about its teratogenic potential and confirmation that it can be transported across the placenta to the fetal compartment [15] as well as its unknown safety for the fetal compartment limit its potential for clinical application [16]. Ferumoxytol, a superparamagnetic iron oxide nanoparticle approved for the clinical treatment of anemia, including during pregnancy, and is a compelling off-label intravascular contrast agent that is increasingly used for MRI [17–20] with regional T1 and T2* shortening, and a long intravascular residency time (∼14.5 hrs). Ferumoxytol has been shown to be well tolerated, associated with no serious adverse events, and implicated in few (1.8% mild and 0.2% moderate) adverse reactions in a multi-center MRI safety study of 4240 subjects stretching over 15 years[21]. Preliminary studies have also suggested no detectable risk for non-human primates [22] and demonstrated safe use in pregnant patients [23].

Current guidelines recommend slow ferumoxytol injections, which limits its ability for dynamic contrast enhanced (DCE MRI), but its intravascular nature is ideal to provide measures of placental blood volume through gestation. Recently, Badachhape et al. estimated fractional blood volume (FBV) in pregnant mice from images acquired before and after contrast injection of intravascular liposomal gadolinium-based contrast agents (GABA) to assess maternal-side placental perfusion and found good agreement with nanoparticle contrast-enhanced computed tomography (CE-CT) [24]. Such blood volume measures can possibly identify placental perfusion deficits that are indicative of local ischemia or fetal growth restriction (FGR). The purpose of this work is to assess the feasibility of ferumoxytol contrast-enhanced MRI to measure maternal placental blood volume in a non-human primate model. This approach was applied in Zika virus-infected and control rhesus macaques [25] at various gestational timepoints.

## Methods

### Animals

This prospective animal study was approved by our institution’s animal care and use committee. Seven pregnant rhesus macaques (maternal body weight 8.20±1.35 kg) were imaged longitudinally at up to 3 timepoints across gestation (days 64.5±1.9, 100.8±3.9, and 145.3±1.8), corresponding to first, second, and third trimester of the rhesus, which has gestation days of 146∼180 full term. Four animals received a Zika virus (ZIKV) injection of the Puerto Rican Zika virus/H.sapiens-tc/PUR/2015/PRVABC59_v3c2 (PR ZIKV) strain into the amniotic fluid: three animals with 10^4^ plaque forming units (PFU) and one animal with 10^5^ PFU. Three control rhesus macaques received a saline injection instead. All injections occurred at gestational age 54.7±1.9 days. Prior to imaging, sedation was completed by injection of up to 10 mg/kg ketamine, followed by intubation and maintenance anesthesia by inhalation of a mixture of oxygen and 1.5% isoflurane.

### MRI Data Acquisition

All scans were performed on a 3.0 T MRI system (Discovery 750, GE Healthcare, Waukesha, WI, USA) using a 32-channel torso coil (Neocoil, Pewaukee, WI, USA) with the animal in the right lateral decubitus position. Blood volume measurements were based on T1 mapping before and after the injection of ferumoxytol. T1 mapping was performed with a respiratory-gated center out, 3D radial, variable flip angle (VFA) spoiled gradient echo (SPGR) sequence[26] (TR = 6.0 ms, TE = 1.2 ms, imaging FOV = 200 x 200 mm^2^, BW = 125 kHz, flip angle = 2°, 6°, 10°, 14°, scan time = 8 min 46 sec). After pre-contrast imaging, Ferumoxytol (4mg/kg with respect to maternal body weight, diluted 5:1 with saline) was intravenously administered with a power injector. Approximately 30 minutes later, the T1 mapping sequence was repeated with identical parameters. Separate to VFA imaging, a vendor supplied 2D Bloch-Siegert sequence was used to perform B1+ mapping. This B1+ map covered the imaging volume of interest contiguously. Figure 1 shows the MRI acquisition workflow.

**Figure 1.**
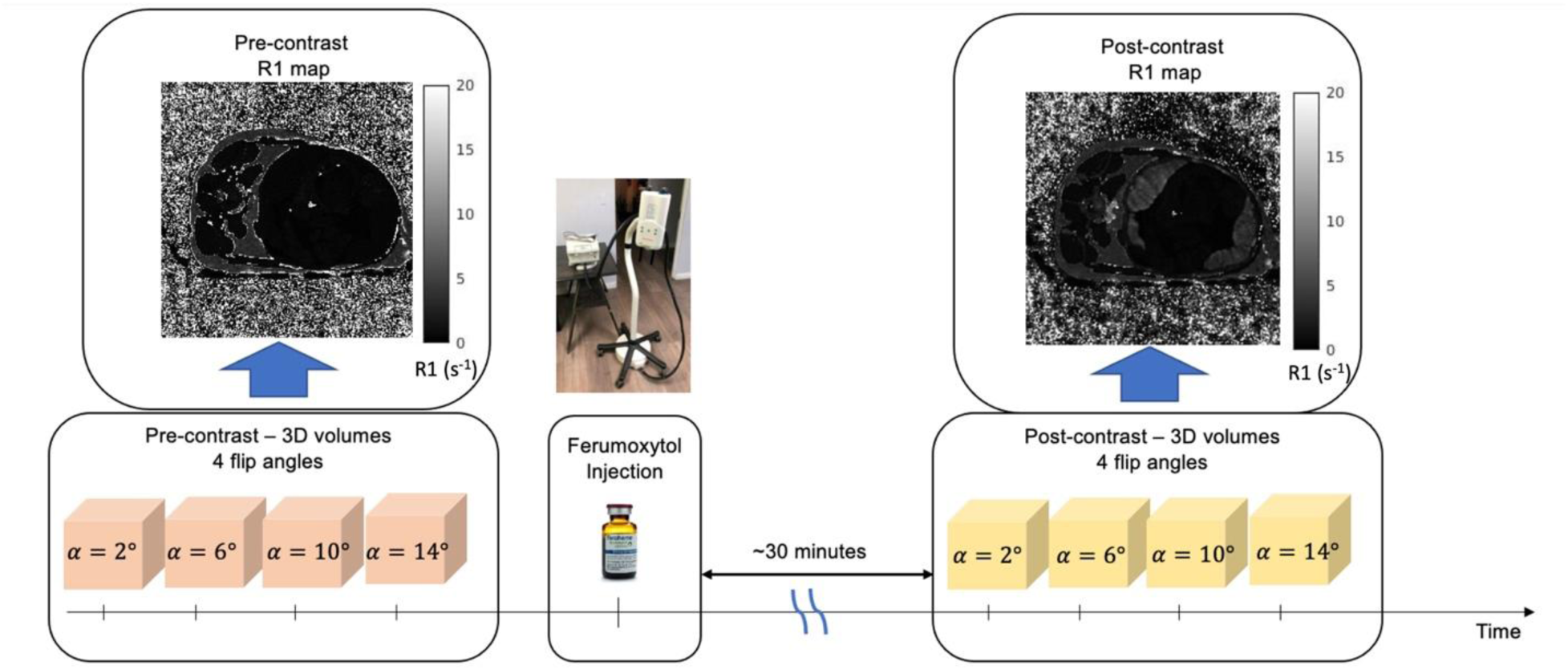
Variable flip angle T1 mapping protocol for Ferumoxytol MRI in rhesus. T1 mapping was performed with a respiratory-gated center out, 3D radial variable flip angle T1 weighted spoiled gradient echo sequence. After pre-contrast image acquisition, ferumoxytol was intravenously administered at 4mg/kg with a power injector. The same T1 mapping sequence was performed approximately 30 minutes after contrast injection. T1 maps were obtained using an in-house toolbox for complex fitting to spoiled gradient echo signals.

### Image Registration and Segmentation

To minimize effects from maternal and fetal motion on blood volume estimates, registration of the pre- and post-contrast R1 maps was performed based on the *α* = 6° T1 weighted scans. Fully automated registration was performed using Advanced Normalization Tools (ANTs) [27] with user-defined symmetric normalization (SyN), including affine followed with deformable transformation, with mutual information as optimization metric. Each registration metric uses a 3-level image pyramid with each level 200 iterations at most. The result of the registration is then applied as a matrix transform to all the other flip angles, prior to T1 fitting. Placental segmentations were conducted manually on the heavily T1-weighted 14° flip angle post-contrast images using an inhouse segmentation tool (MATLAB). In addition, a circular ROI was drawn on the uterine artery to measure R1 values in blood.

### VFA T1-Mapping Workflow[28]

Pixel-wise T1 values were determined via fitting of the four flip angles with the corresponding T1 and signal density in a SPGR signal model. B1+ inhomogeneities were accounted for by using B1+ maps, which were re-sampled using bi-linear interpolation to match the dimension and voxel grid of the VFA images. Signal fitting was performed in the complex domain with mean square error minimization between the predicted signal and the measurement data using an in an in-house tool written in C++.

### Blood volume calculation

Maternal placental blood volumes were estimated using the pre- and post-contrast R1 (1/T1) maps generated by variable flip angle fitting. ΔR1 maps were obtained by pixel-wise subtraction of the two registered R1 maps. Three quantitative blood volume measurements were derived:

1. **<u>maternal fractional blood volume (mFBV),</u>** which is the fractional amount of placental volume occupied by maternal blood, calculated as the ratio between median ΔR1 in placenta, DeltaR1_palcenta) and maternal blood, DeltaR1_vessel, hwere assessed in the uterine artery:

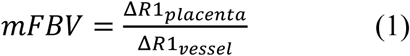 Regional mFBV maps were calculated as the ratio of pixel-wise ΔR1 in placenta and median Δ*R1* of blood. Placental regions containing mFBV value of 1 suggests 100% blood content (or 0% tissue) in that region.
2. **<u>Maternal placental blood volume (mPBV</u>**), which is the total blood volume (in milliliters) in the placenta due to maternal contribution. This is calculated as placental volume, *V*_placenta_, multiplied by mean mFBV.

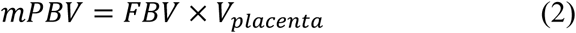
3. **<u>Total maternal blood volume (mTBV</u>**), which is the total blood volume (in milliliters) in the rhesus macaque animals, not counting the fetal blood. This is calculated as a ratio of amount of ferumoxytol (mol) in placental tissue over concentration of ferumoxyol (M) in blood *C*_*b_total*_.

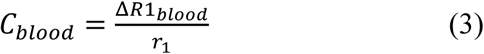

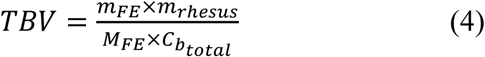 We assume a fast-exchange limit of a two-compartment water exchange model since the water protons diffuse quickly across the vessel walls due to the highly permeable nature of the placenta[29]. Therefore, a linear relationship between Ferumoxytol concentration and relaxivity (r_1_=8.6725s^−1^mM^−1^)[30] is assumed.

A paired T-test was used to determine statistical significance in blood volume measurements between controls and Zika-infected rhesus macaques.

## Results

While all ZYKV treated dams experienced a productive infection, the post-delivery analysis showed no significant fetal anomalies and no evidence of vertical transmission of the virus. Histopathological analysis showed varying degrees of presence of pathology in both groups, virus-exposed animals and controls, with no statistically significant difference in pathology scores.

Measurements of T1/R1 values and blood volume were successfully achieved for all scans. Figure 2 shows a representative example from a control animal (#3): pre- and post-contrast anatomical T1-weighted scans (left to right: 2°, 6°, 10°, 14°), as well as the corresponding R1 maps on the right.

**Figure 2.**
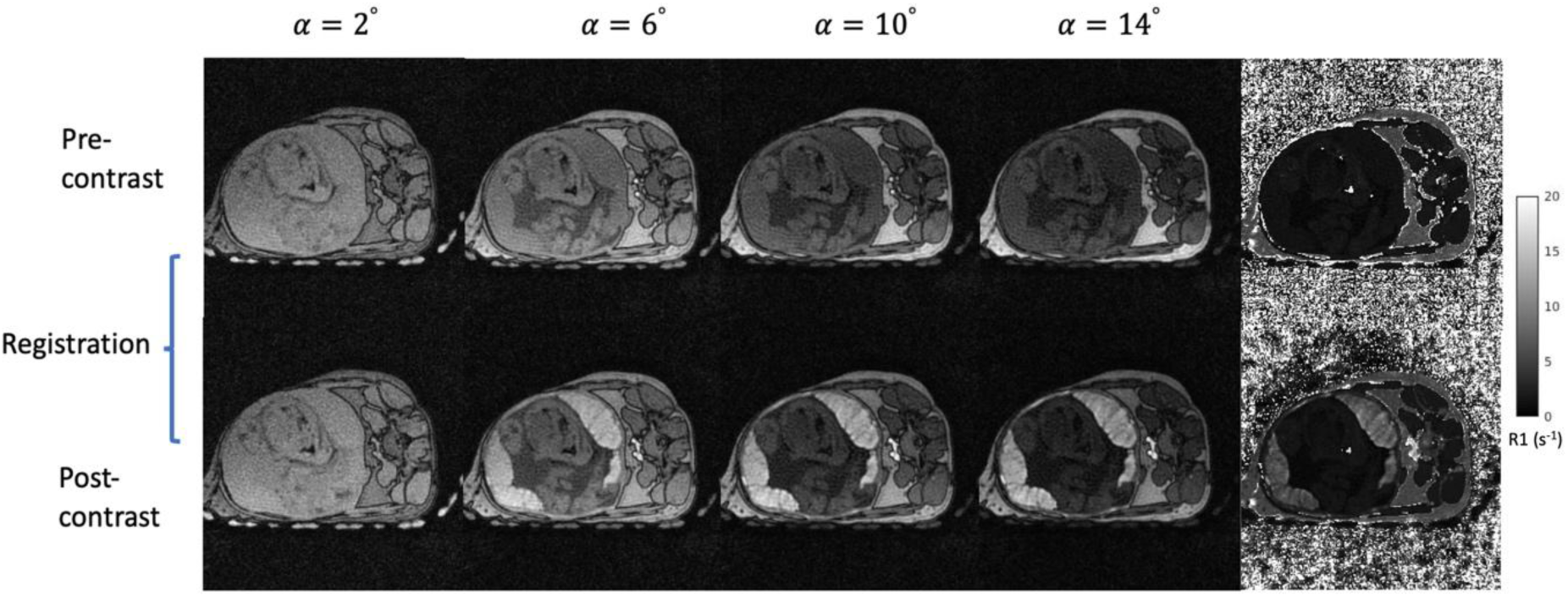
Variable flip angle images. Left four columns show representative Pre- and post-contrast (top and bottom) T1-weighted images at each flip angle (left to right: 2°, 6°, 10°, 14°) of control animal #3. The R1 fitting result is shown on the right. Pre- and post-contrast T1 weighted images were registered using affine then non-rigid registration.

Figure 3 shows 6 non-consecutive mFBV maps (values are color-coded with a heat map scale) of the same animal as that of Figure 3, overlaid on the corresponding T1-weighted SPGR anatomical images obtained for a flip angle of 14°. mFBV range from 0 to 1, with 1 meaning the region has 100% blood (such as it being a blood vessel). mFBV exhibits a regionally heterogeneous distribution both across slices and within each slice. The white arrows indicate regions with high blood volume, which could correspond to spiral artery inflow zones of the placenta.

**Figure 3.**
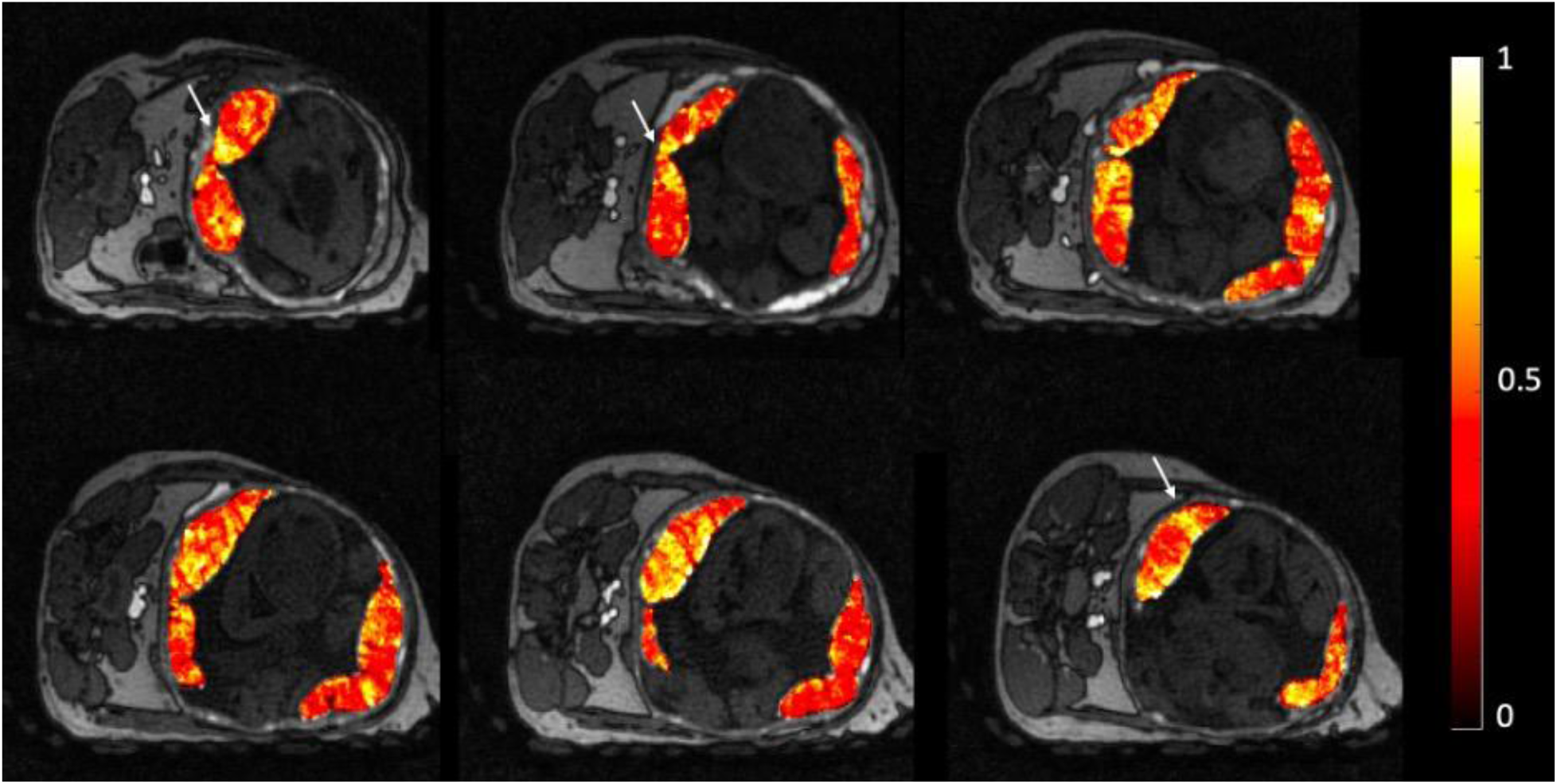
Representative fractional blood volume maps. Six selected slices (out of ∼150 slices covering the placenta) of maternal fractional blood volume maps are overlaid on top of the corresponding anatomical T1-weighted images (animal # 2, gestation day = 146). A value of 1 indicates a region that contains 100% blood. The maternal fractional blood volume (mFBV) exhibits a regionally strongly heterogeneous distribution with a mean of 0.48 in this case. White arrows point at regions with high maternal blood volume, presumably spiral artery inflow zones.

Table 1 shows mFBV, mPBV, and Δ*R*1_*blood*_ values for each animal at each gestation time point scanned and Figure 4 shows median mFBV and mPBV measurements for each animal across gestation ages. The data show similar trends across the cohort: maternal fractional blood volume measurements stay fairly flat with only small increases or decreases while maternal placental blood volumes increase steeply, driven by the volume increases of the placenta.

**Figure 4.**
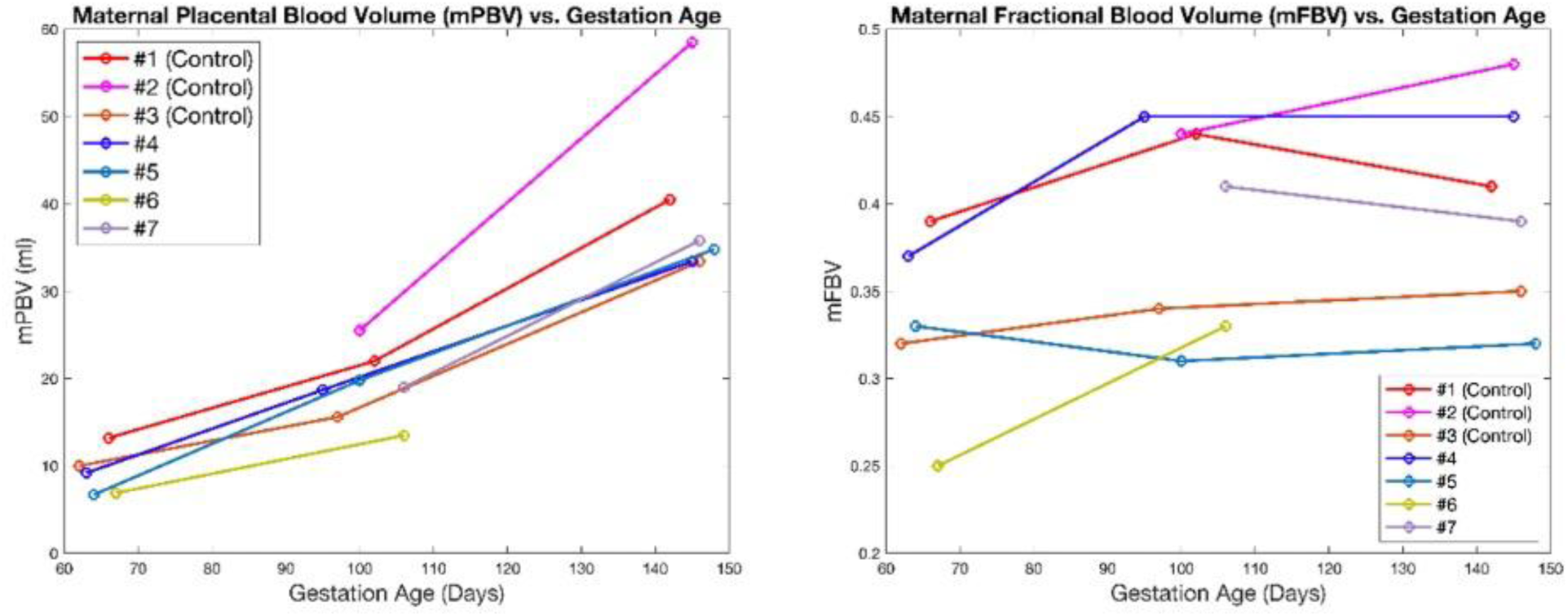
Longitudinal plots of maternal placental blood volume. This shows consistent increase in blood volume with respect to gestation age. The maternal fractional blood volumes show a more heterogeneous distribution: most animals tend to have an increase in mean blood volume with gestation, whereas two animals show decrease in blood volume from the second to the third gestational timepoint. Note that the zika infection in this study resulted in very modest pathology difference, which is supported by our data showing no significant difference in blood volume between the two treatment groups.

**Table 1.**
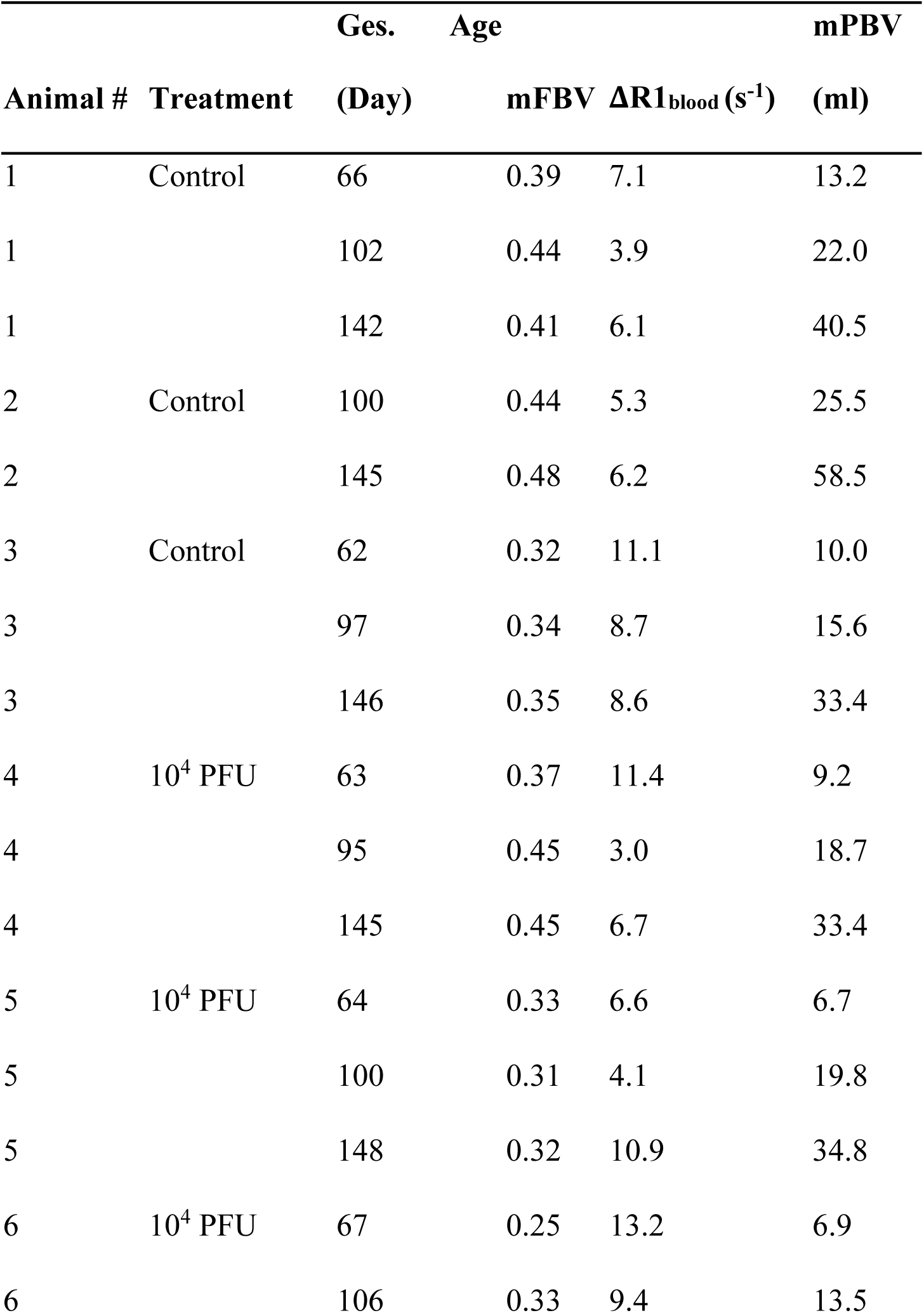
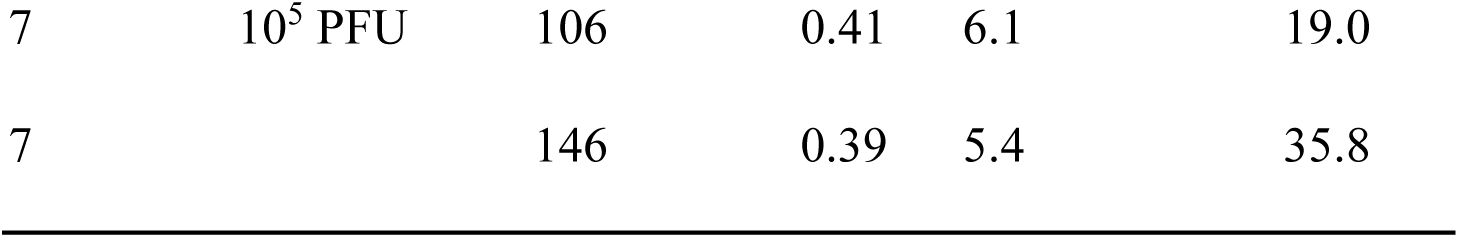
Rhesus blood volume measurement results. This table shows maternal fractional blood volume (mFBV), maternal placental blood volume (mPBV), and pre- and post-contrast R1 difference in blood vessels (ΔR1_blood_) values for each animal at each gestation time point scanned.

Statistical analyses show no significant differences in mFBV between controls and zika-infected rhesus macaques, which corresponds to our findings from the placental pathology analysis. Hence we are reporting results across the entire cohort and do not distinguish between ZYKA and controls. Average maternal fractional blood volume (mFBV) values across animals show a modest increase with gestational age: 0.33±0.05, 0.38±0.06, and 0.40±0.06 for gestational time points (GTPs) 1, 2, and 3 respectively. Meanwhile, average mPBV was measured as 11.1±5.0 ml, 19.2±4.0 ml, and 39.4±9.7 ml for GTPs 1, 2, and 3 respectively, demonstrating a consistentn and substantial increases with gestation age.

Figure 5 shows histograms of maternal fractional blood volume, ΔR1 in placenta, and ΔR1 in blood averaged over the 4 animals imaged for all three gestational time points. Median TBV across all scans is 740.3 ml, and median TBV normalized by maternal body weight is 93.4 ml/kg.

**Figure 5.**
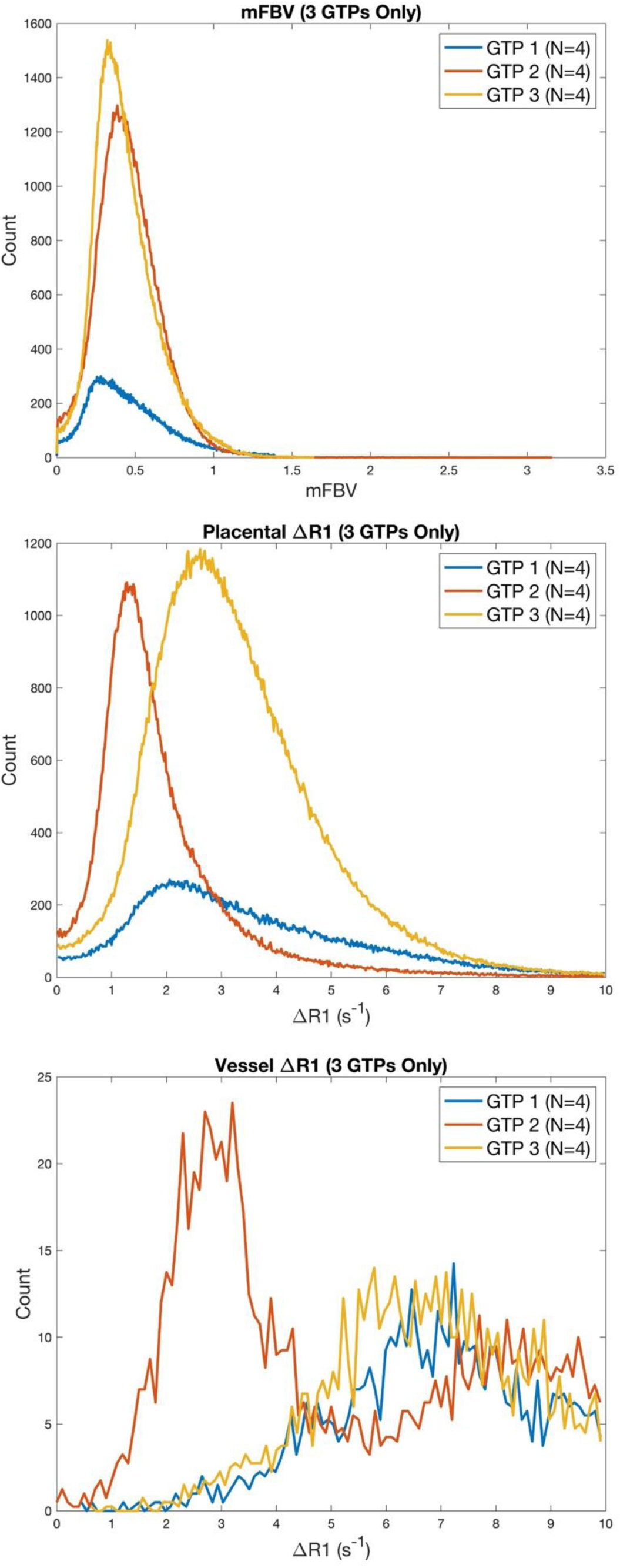
Pixel-wise blood volume histograms. This shows (top to bottom) maternal fractional blood volume, ΔR1 in placenta, and ΔR1 in blood averaged over all animals imaged for all three gestational time points (GTPs).

## Discussion

This pilot study demonstrated the feasibility of quantitatively measuring maternal placental blood volume using ferumoxytol enhanced MRI. Results in rhesus macaques demonstrated a high degree of success across animals, with excellent image quality. Normalization of the R1 mapping by the R1 increase in blood improved quantitative agreement across animals. The maternal blood volume fraction of the placenta was relatively stable across animals and throughout gestation, with an overall increase in placenta volume due to volumetric growth of the placenta. Overall, this study establishes ferumoxytol blood volume mapping as a promising technique to quantify placental vascular function in animal models and in human studies.

Ferumoxytol increased R1 relaxivity in both placental tissue and in blood vessels, but no enhancement in signal was observed quantitatively or visually in fetal tissue. This agrees with previous study in humans that were limited to visually evaluated images[31]. This continues to provide additional evidence supporting the used of ferumoxytol given that it does not cross the maternal-fetal barrier. This in contrast to current Gd based agents, which do pass to fetus in trace amounts. However, this also means that ferumoxytol blood volume measures only represent maternal blood volume and these measures do not inform on potential abnormalities in the fetal circulation. Although clinical literature suggests maternal blood-flow deficit in common pregnancy disorders, studies are needed to verify the utility of maternal only blood volume measures using contrast enhanced MRI in identifying pathology.

The mean mFBV across all animals increased with gestation age, although each animal showed slightly different trends individually. This slight increase agrees with the findings by Badachhape et al. in a pregnant mouse model[24]. This suggests possible advances in placental vascular development with gestation. Mean mPBV also showed significant increases with gestation age for all animals, especially from the second to third trimester gestational time point. This is largely due to the overall growth in the placenta volume over gestation, coupled with the slight increase in mFBV.

The mean TBV measured in this study from all scans is 50.4% higher than previous TBV measurement in non-pregnant adult rhesus macaques of 62.1 ml/kg using an indicator dilution methodology[32]. This is within the expected range of normal percent of blood volume increase during human pregnancy[33]. However, some of the observed TBV variations are not physiological plausible. Our TBV measures are dependent on precise knowledge of the amount of ferumoxytol injected and assumes an exact dose of ferumoxytol is delivered to the animal. Errors in dosing may proportionately propagate to errors in dosing given. Animals in this study received a bolus delivery of ferumoytol for dynamic contrast enhanced (DCE) MRI [22]. We suspect that imperfect mixing with saline, incomplete injections solution in line, or extravasation might have occurred in some instances and corrupted TBV estimates. Such situations are avoidable in pharmacy-prepared larger volumes generally used in slow infusion treatments in human investigations, but with the limitations associated with slow infusion rather than concentrated rapid bolus injections. Future studies should be aware of this potential limitation and should use normalized measures (mFBV, mPBV) whenever possible.

Local analysis of mFBV showed a heterogeneous distribution of high blood volume zones (∼100% FBV), which could correspond to inflow vascular regions for placental functional units. The histogram analysis shows higher skewness for both mFBV and Δ*R*1 for the first time point. This could be due to less concentrated central perfusion regions at early gestation[34]. We see a left shift in ΔR1 values for both placental and blood regions in the second gestational time point, which could be due to imperfect contrast injection, since our power injector was normally used for larger volume and higher concentration injections for humans. However, this effect is does not affect our mFBV measurements since they do not require prior knowledge of the amount of contrast injected.

There are a few limitations of our study. Maternal and fetal motion during scan can result in motion artifact and imperfect registration between pre- and post-contrast images. Since our animals were sedated for the scans, fetal motion was the main issue since it warps the placental shapes. This occurs more often at higher gestational ages. One animal (#8) was excluded from analysis due to significant fetal motion during the time between pre- and post-contrast scans. Additionally, the limited number of animals and scan timepoints limits the extent of statistical analysis of our data. Lastly, the ZIKV infection model has previously been shown to have very modest placental pathology in a related study[25]. The modest pathological outcomes of the placentas may result in small blood volume value differences between the control animals and the Zika-infected animals, which then could not serve as a direct indication of clinical detectability of pathology using blood volume measurements.

The use of ferumoxytol contrast agent and a non-human primate model provides insight on the translational potential of in-vivo blood volume measurements. Future work includes blood volume measurements on Rhesus animals with induced pathology (e.g. regional placental infarction) to directly visualize pathology detectability. Additionally, the clinical use of Ferumoxytol allows for quantitative blood volume measurement and analysis on humans with fetal growth restrictions.

## Data Availability

All data produced in the present study are available upon reasonable request to the authors.

## Acknowledgements

We gratefully acknowledge GE Healthcare for research support of UW-Madison, and AMAG Pharmaceuticals for providing ferumoxytol used in our imaging procedures. We also thank the Wisconsin National Primate Research Center (WNPRC) Veterinary, Scientific Protocol Implementation, and Animal Services staff for providing animal care, and assisting in procedures including breeding, pregnancy monitoring, and sample collection. Lastly, this work was supported by the National Institutes of Health (grants R01 HD103443, R21 AI129308, U01 HD087216, and T32 HD041921).

## References

[1] N. Siauve, G.E. Chalouhi, B. Deloison, M. Alison, O. Clement, Y. Ville, L.J. Salomon, Functional imaging of the human placenta with magnetic resonance., Am J Obstet Gynecol 213 (2015) S103–14. 10.1016/j.ajog.2015.06.045.

[2] J.M. Roberts, C. Escudero, The placenta in preeclampsia., Pregnancy Hypertens 2 (2012) 72–83. 10.1016/j.preghy.2012.01.001.

[3] I. Brosens, R. Pijnenborg, L. Vercruysse, R. Romero, The “Great Obstetrical Syndromes” are associated with disorders of deep placentation., Am J Obstet Gynecol 204 (2011) 193–201. 10.1016/j.ajog.2010.08.009.

[4] S. Hong, Y. Le, K.U. Lio, T. Zhang, Y. Zhang, N. Zhang, Performance comparison of ultrasonography and magnetic resonance imaging in their diagnostic accuracy of placenta accreta spectrum disorders: a systematic review and meta-analysis, Insights Imaging 13 (2022) 50. 10.1186/s13244-022-01192-w.

[5] J.A. Macdonald, P.A. Corrado, S.M. Nguyen, K.M. Johnson, C.J. Francois, R.R. Magness, D.M. Shah, T.G. Golos, O. Wieben, Uteroplacental and Fetal 4D Flow MRI in the Pregnant Rhesus Macaque., J Magn Reson Imaging 49 (2019) 534–545. 10.1002/jmri.26206.

[6] K.L. Thornburg, N. Marshall, The placenta is the center of the chronic disease universe., Am J Obstet Gynecol 213 (2015) S14–20. 10.1016/j.ajog.2015.08.030.

[7] D.J. Barker, The fetal and infant origins of adult disease., BMJ 301 (1990) 1111. 10.1136/bmj.301.6761.1111.

[8] Society for Maternal-Fetal Medicine Publications Committee, E. Berkley, S.P. Chauhan, A. Abuhamad, Doppler assessment of the fetus with intrauterine growth restriction., Am J Obstet Gynecol 206 (2012) 300–8. 10.1016/j.ajog.2012.01.022.

[9] K.D. Ludwig, S.B. Fain, S.M. Nguyen, T.G. Golos, S.B. Reeder, I.M. Bird, D.M. Shah, O.E. Wieben, K.M. Johnson, Perfusion of the placenta assessed using arterial spin labeling and ferumoxytol dynamic contrast enhanced magnetic resonance imaging in the rhesus macaque., Magn Reson Med 81 (2019) 1964–1978. 10.1002/mrm.27548.

[10] I. Huen, D.M. Morris, C. Wright, G.J.M. Parker, C.P. Sibley, E.D. Johnstone, J.H. Naish, R1 and R2* changes in the human placenta in response to maternal oxygen challenge, Magn Reson Med 70 (2013) 1427–1433. 10.1002/mrm.24581.

[11] A.E.P. Ho, J. Hutter, L.H. Jackson, P.T. Seed, L. McCabe, M. Al-Adnani, A. Marnerides, S. George, L. Story, J. V. Hajnal, M.A. Rutherford, L.C. Chappell, T2∗ Placental Magnetic Resonance Imaging in Preterm Preeclampsia: An Observational Cohort Study, Hypertension 75 (2020) 1523–1531. 10.1161/HYPERTENSIONAHA.120.14701.

[12] D. Liu, X. Shao, A. Danyalov, T. Chanlaw, R. Masamed, D.J.J. Wang, C. Janzen, S.U. Devaskar, K. Sung, Human Placenta Blood Flow During Early Gestation With Pseudocontinuous Arterial Spin Labeling MRI., J Magn Reson Imaging 51 (2020) 1247–1257. 10.1002/jmri.26944.

[13] K.Y. Oh, V.H.J. Roberts, M.C. Schabel, K.L. Grove, M. Woods, A.E. Frias, Gadolinium Chelate Contrast Material in Pregnancy: Fetal Biodistribution in the Nonhuman Primate., Radiology 276 (2015) 110–8. 10.1148/radiol.15141488.

[14] Y.O. Tanaka, S. Sohda, S. Shigemitsu, M. Niitsu, Y. Itai, High temporal resolution dynamic contrast MRI in a high risk group for placenta accreta., Magn Reson Imaging 19 (2001) 635–42. 10.1016/s0730-725x(01)00388-5.

[15] J.O. Lo, V.H.J. Roberts, M.C. Schabel, X. Wang, T.K. Morgan, Z. Liu, C. Studholme, C.D. Kroenke, A.E. Frias, Novel Detection of Placental Insufficiency by Magnetic Resonance Imaging in the Nonhuman Primate, Reproductive Sciences 25 (2018) 64–73. 10.1177/1933719117699704.

[16] F. Garcia-Bournissen, A. Shrim, G. Koren, Safety of gadolinium during pregnancy., Can Fam Physician 52 (2006) 309–10.

[17] M.R. Bashir, L. Bhatti, D. Marin, R.C. Nelson, Emerging applications for ferumoxytol as a contrast agent in MRI., J Magn Reson Imaging 41 (2015) 884–98. 10.1002/jmri.24691.

[18] S.S. Vasanawala, K.-L. Nguyen, M.D. Hope, M.D. Bridges, T.A. Hope, S.B. Reeder, M.R. Bashir, Safety and technique of ferumoxytol administration for MRI., Magn Reson Med 75 (2016) 2107–11. 10.1002/mrm.26151.

[19] M.D. Hope, T.A. Hope, C. Zhu, F. Faraji, H. Haraldsson, K.G. Ordovas, D. Saloner, Vascular Imaging With Ferumoxytol as a Contrast Agent., AJR Am J Roentgenol 205 (2015) W366–73. 10.2214/AJR.15.14534.

[20] G.B. Toth, C.G. Varallyay, A. Horvath, M.R. Bashir, P.L. Choyke, H.E. Daldrup-Link, E. Dosa, J.P. Finn, S. Gahramanov, M. Harisinghani, I. Macdougall, A. Neuwelt, S.S. Vasanawala, P. Ambady, R. Barajas, J.S. Cetas, J. Ciporen, T.J. DeLoughery, N.D. Doolittle, R. Fu, J. Grinstead, A.R. Guimaraes, B.E. Hamilton, X. Li, H.L. McConnell, L.L. Muldoon, G. Nesbit, J.P. Netto, D. Petterson, W.D. Rooney, D. Schwartz, L. Szidonya, E.A. Neuwelt, Current and potential imaging applications of ferumoxytol for magnetic resonance imaging., Kidney Int 92 (2017) 47–66. 10.1016/j.kint.2016.12.037.

[21] K.-L. Nguyen, T. Yoshida, N. Kathuria-Prakash, I.H. Zaki, C.G. Varallyay, S.I. Semple, R. Saouaf, C.K. Rigsby, S. Stoumpos, K.K. Whitehead, L.M. Griffin, D. Saloner, M.D. Hope, M.R. Prince, M.A. Fogel, M.L. Schiebler, G.H. Roditi, A. Radjenovic, D.E. Newby, E.A. Neuwelt, M.R. Bashir, P. Hu, J.P. Finn, Multicenter Safety and Practice for Off-Label Diagnostic Use of Ferumoxytol in MRI., Radiology 293 (2019) 554–564. 10.1148/radiol.2019190477.

[22] S.M. Nguyen, G.J. Wiepz, M. Schotzko, H.A. Simmons, A. Mejia, K.D. Ludwig, A. Zhu, K. Brunner, D. Hernando, S.B. Reeder, O. Wieben, K. Johnson, D. Shah, T.G. Golos, Impact of ferumoxytol magnetic resonance imaging on the rhesus macaque maternal-fetal interface†., Biol Reprod 102 (2020) 434–444. 10.1093/biolre/ioz181.

[23] M.A. Kliewer, C.G. Bockoven, S.B. Reeder, A.R. Bagley, E.A. Sadowski, J.I. Iruretagoyena, M.J. Beninati, M.K. Fritsch, Ferumoxytol-enhanced MR demonstration of changes to internal placental structure in placenta accreta spectrum: Preliminary findings., Placenta 134 (2023) 1–8. 10.1016/j.placenta.2023.02.003.

[24] A.A. Badachhape, L. Devkota, I. V Stupin, P. Sarkar, M. Srivastava, E.A. Tanifum, K.A. Fox, C. Yallampalli, A. V Annapragada, K.B. Ghaghada, Nanoparticle Contrast-enhanced T1-Mapping Enables Estimation of Placental Fractional Blood Volume in a Pregnant Mouse Model., Sci Rep 9 (2019) 18707. 10.1038/s41598-019-55019-8.

[25] D.P. Seiter, S.M. Nguyen, T.K. Morgan, L. Mao, D.M. Dudley, D.H. O’connor, M.E. Murphy, K.D. Ludwig, R. Chen, A. Dhyani, A. Zhu, M.L. Schotzko, K.G. Brunner, D.M. Shah, K.M. Johnson, T.G. Golos, O. Wieben, Ferumoxytol Dynamic Contrast Enhanced Magnetic Resonance Imaging Identifies Altered Placental Cotyledon Perfusion in Rhesus Macaques., Biol Reprod (2022). 10.1093/biolre/ioac168.

[26] S.C.L. Deoni, Quantitative relaxometry of the brain., Top Magn Reson Imaging 21 (2010) 101–13. 10.1097/RMR.0b013e31821e56d8.

[27] B.B. Avants, N.J. Tustison, M. Stauffer, G. Song, B. Wu, J.C. Gee, The Insight ToolKit image registration framework., Front Neuroinform 8 (2014) 44. 10.3389/fninf.2014.00044.

[28] R. Heule, C. Ganter, O. Bieri, Variable flip angle T1 mapping in the human brain with reduced T2 sensitivity using fast radiofrequency-spoiled gradient echo imaging., Magn Reson Med 75 (2016) 1413–22. 10.1002/mrm.25668.

[29] C.M. Colbert, M.A. Thomas, R. Yan, A. Radjenovic, J.P. Finn, P. Hu, K.-L. Nguyen, Estimation of fractional myocardial blood volume and water exchange using ferumoxytol-enhanced magnetic resonance imaging., J Magn Reson Imaging 53 (2021) 1699–1709. 10.1002/jmri.27494.

[30] G. Knobloch, T. Colgan, C.N. Wiens, X. Wang, T. Schubert, D. Hernando, S.D. Sharma, S.B. Reeder, Relaxivity of Ferumoxytol at 1.5 T and 3.0 T., Invest Radiol 53 (2018) 257–263. 10.1097/RLI.0000000000000434.

[31] M.A. Kliewer, C.G. Bockoven, S.B. Reeder, A.R. Bagley, M.K. Fritsch, Ferumoxytol-enhanced magnetic resonance imaging with volume rendering: A new approach for the depiction of internal placental structure in vivo, Placenta 131 (2023). 10.1016/j.placenta.2022.12.001.

[32] T.R. Hobbs, S.W. Blue, B.S. Park, J.J. Greisel, P.M. Conn, F.K.-Y. Pau, Measurement of Blood Volume in Adult Rhesus Macaques (Macaca mulatta)., J Am Assoc Lab Anim Sci 54 (2015) 687–93.

[33] F. Hytten, Blood volume changes in normal pregnancy., Clin Haematol 14 (1985) 601–12.

[34] J. Hutter, P.J. Slator, L. Jackson, A.D.S. Gomes, A. Ho, L. Story, J. O’Muircheartaigh, R.P.A.G. Teixeira, L.C. Chappell, D.C. Alexander, M.A. Rutherford, J. V. Hajnal, Multi-modal functional MRI to explore placental function over gestation, Magn Reson Med 81 (2019) 1191–1204. 10.1002/mrm.27447.

